# Role of Weather Factors in COVID-19 Death Growth Rates in Tropical Climate: A Data-Driven Study Focused on Brazil Manuscript

**DOI:** 10.1101/2020.09.13.20193532

**Authors:** Rahul Kalippurayil Moozhipurath

## Abstract

**Background:** Brazil reported 123,780 deaths across 27 administrative regions, making it the second-worst affected country after the US in terms of COVID-19 deaths as of 3 September 2020. Understanding the role of weather factors in COVID-19 in Brazil is helpful in the long-term mitigation strategy of COVID-19 in other tropical countries because Brazil experienced early large-scale outbreak among tropical countries. Recent COVID-19 studies indicate that relevant weather factors such as temperature, humidity, UV Index (UVI), precipitation, ozone, pollution and cloud cover may influence the spread of COVID-19. Yet, the magnitude and direction of those associations remain inconclusive. Furthermore, there is only limited research exploring the impact of these weather factors in a tropical country like Brazil. In this observational study, we outline the roles of 7 relevant weather factors including temperature, humidity, UVI, precipitation, ozone, pollution (visibility) and cloud cover in COVID-19 death growth rates in Brazil.

**Methods:** We use a log-linear fixed-effects model to a panel dataset of 27 administrative regions in Brazil across 182 days (n=3882) and analyze the role of relevant weather factors by using daily cumulative COVID-19 deaths in Brazil as the dependent variable. We carry out robustness checks using case-fatality-rate (CFR) as the dependent variable.

**Findings:** We control for all region-specific time-fixed factors as well as potentially confounding time-varying factors. We observe a significant negative association of COVID-19 daily deaths growth rate in Brazil with weather factors - UVI, temperature, ozone and cloud cover. Specifically, a unit increase in UVI, maximum temperature, and ozone level independently associate with 6.0 percentage points [p<0.001], 1.8 percentage points [p<0.01] and 0.3 percentage points [p<0. 1] decline in COVID-19 deaths growth rate. Further, a percentage point increase in cloud cover associates with a decline of 0.148 percentage points [p<0.05] in COVID-19 deaths growth rate. Surprisingly, contrary to other studies, we do not find evidence of any association between COVID-19 daily deaths growth rate and humidity, visibility and precipitation. We find our results to be consistent even when we use the CFR as the dependent variable.

**Interpretation:** We find independent protective roles of UVI, temperature, ozone and cloud cover in mitigating COVID-19 deaths growth rate, even in a tropical country like Brazil. We observe these results to be consistent across various model specifications, especially for UVI and cloud cover, even after incorporating additional time-varying weather parameters such as dewpoint, pressure, wind speed and wind gust. These results could guide health-related policy decision making in Brazil as well as similar tropical countries.

## 1 Introduction

Brazil reported 123,780 deaths across 27 administrative regions, making it the second-most affected country after the US in terms of COVID-19 deaths as of 3 September 2020^1^. Understanding the role of weather factors in COVID-19 in Brazil would be helpful in the long-term mitigation strategy of COVID-19 in other tropical countries because Brazil experienced early large-scale outbreak among tropical countries. Recent COVID-19 studies indicate that relevant weather factors such as temperature, humidity, UV Index (UVI), precipitation, ozone, pollution and cloud cover may influence the spread of COVID-19. Yet, the magnitude and direction of those associations in general and also specifically for tropical countries remain inconclusive. A majority of the studies considered only a subset of these weather factors and they did not take into account various region-specific time-constant factors as well as the potentially lagged effects of the weather factors, which might confound the results.

So far, studies looking at the impact of these weather factors on COVID-19 in a tropical country like Brazil are limited. Brazil is an interesting country for studying the impact of weather parameters on COVID-19 primarily due to three reasons. Firstly, Brazil is the most affected country in tropics and second most affected country in the world after the US in terms of COVID-19 deaths. Secondly, Brazil is the fifth largest country with a majority of its territory in Southern Hemisphere (93% of the landmass in Southern Hemisphere^2^) – spanning across 5°N and 33°S. Initial stages of COVID-19 started in Brazil during their autumn (Southern Hemisphere), and subsequently, Brazil transitioned from autumn to winter. The dynamics of COVID-19 in autumn to winter transition may improve our understanding of the role of weather factors, especially for similar regions in the northern hemisphere, when they transition from autumn to winter. Thirdly, a large area of Brazil’s geography (81.4%)^3^ is tropical, experiencing a wide range of seasons and weather conditions prominent in a tropical climate, enabling us to draw early conclusions on how weather factors might impact COVID-19 in tropical areas^4^. Therefore, the results of such a study in Brazil can guide further health-related policy decision making in similar tropical countries.

To the best of our knowledge, so far, no empirical study has comprehensively explored the role of these different relevant weather factors in COVID-19 death growth rates covering 27 different administrative regions in Brazil, controlling for various other time-varying factors well as region-specific time-constant factors. In this observational study, we empirically describe the role of these weather factors in COVID-19 death grow rates in Brazil. We observe a significant negative association of daily growth rate of cumulative COVID-19 deaths (COVID-19 daily death growth rate) in Brazil with weather factors - UVI, temperature, ozone and cloud cover, indicting their respective protective roles. Surprisingly, contrary to other studies, we do not find any association between COVID-19 daily death growth rates and humidity, visibility and precipitation. We find our results to be consistent even when we use the case-fatality-rate (CFR) as the dependent variable.

## 2 Role of Relevant Weather Factors in COVID-19

Studies indicate that the impact of weather factors on respiratory viral infections and deaths comes from three major mechanisms^5^. Firstly, weather factors affect the survival and transmission of viruses^5^. Respiratory viral infection occurs primarily through contact, droplet spray, aerosol or fomite or surface transmission^5^ and weather factors may play a role in this transmission^5^. Secondly, weather factors play a role in human behaviour affecting the likelihood of being in contact with an infected person and thereby the transmission of viruses^5^. Thirdly, weather factors such as UVB Radiation can modulate the immune responses via its role in vitamin D synthesis^5^.

Prior studies show that weather factors^6^ such as humidity^7^, UVI^3^, temperature^8^ and precipitation^8,9^ play a substantial role in the transmission of viruses such as influenza. Early studies indicate that weather factors such as temperature^10^ and humidity^10,11^ may influence the spread of COVID-19 and plausibly deaths. Temperature and humidity primarily impact the survival period of the virus on different surfaces influencing its transmission^12–14^. Thus, their protective role may be due to their impact on the viability of viruses as they affect the viral membranes^5^. On the contrary, temperature and humidity also influence the behaviour of people. For example, a higher temperature may also increase their likelihood of outdoor activities and thereby increasing the possibility of transmission.

Moisture-based weather factors like precipitation may play a role in converting dry environment into moist and cold conditions, making it more conducive for the survival and transmission of viruses^15^. However, precipitation and cloud cover also influence the behaviour of people, limiting their likelihood of outdoor activities and thereby potentially impacting the transmission of COVID-19.

Further, air pollution^16^ may be a significant risk factor for respiratory infection as it affects the immune system^17^, as well as denser particulate matter, could carry coronavirus for more extended periods as well as across larger distances^17^. Studies also indicate a significant association of air pollution and COVID-19 death rate^17^. Furthermore, studies show that the presence of air pollutants like ozone is also associated with an increased risk of ARDS (Acute Respiratory Distress Syndrome)^18^, which is a significant risk factor in COVID-19. On the contrary, some studies also indicate a protective role of ozone as it reduces the transmission of viral diseases like influenza^19^ and plausibly COVID-19^20^, plausibly due to anti-viral effects^20^.

UV Radiation, another weather factor, plays a protective role in two ways. UV Radiation inactivates viruses in fomite transmission, thereby mitigating the viral spread and deaths^21^. UV Radiation, specifically UVB also plays a significant role in vitamin D skin synthesis^22–26^. Even in a sunny and tropical country like Brazil, studies indicate that Vitamin D deficiency is common^27^. UVB Radiation and the likelihood of skin exposure & skin synthesis vary substantially several factors such as seasons^28^, time^28^, latitude^28^, altitude^28^, active lifestyle^29,30^, dietary habits^31^,^28^, food fortification^28^, age^28^ and skin colour^28^. UV index measures the intensity of UV radiation and is used as a warning for sunburn risk, mostly by solar UVB radiation spectrum 290-315 nm^32^, the spectrum which helps in vitamin D skin synthesis^33^. Early studies show a protective role of UVB and vitamin D in COVID-19^34,35,36,37,38^. 1,25-dihydroxyvitamin D [1,25 (OH)_2_D], an active vitamin D form, plays an essential role in modulating innate and adaptive immune systems^39,40^, renin-angiotensin system (RAS)^40–42^ as well as in modulating the inflammatory response ^39,40^. Emerging COVID-19 studies indicate that vitamin D deficiency might be a risk factor not only for incidence^36,37^, but also for severity^43 44^and mortality^38,45,46^. Therefore, based on the prior studies, we consider temperature, humidity, UV Index, precipitation, ozone, visibility and cloud cover to be the relevant factors to consider for exploring the role of weather in COVID-19 deaths growth rate.

## 3 Evaluation of Previous Studies on Weather Factors & COVID-19

We anticipate that studies exploring the role of weather in COVID-19 need to consider all the above relevant weather factors, as considering only a subset of these factors may confound and limit the reliability of results^47^. Early studies also indicate that a tropical country like Brazil shows some substantial variation from higher latitude countries with low temperature and low UV Index or those with periods under winter^4^. We argue that conclusions of studies with over-representation of locations with a lower temperature or lower UV^47^ either due to the study period or due to specific regions may have limited applicability in a tropical country like Brazil. Furthermore, prior studies indicate that an extended study period helps in strengthening the evidence for the association between these factors and COVID-19^4^. Therefore, it is vital to consider relevant weather parameters as well as the regional characteristics and the duration of the study.

Studies also indicate the importance of considering lags, as there is usually a substantial delay in the onset of symptoms, testing, hospitalisation and deaths^47,48^. Furthermore, humans can store vitamin D produced via the UV Index to be used later^34^. According to prior studies, time-constant factors such as population, genetic profile, culture, density, age structure, gender, underlying disease, co-morbidities, skin pigmentation, gender, mobility, underlying conditions, diet or socio-economic factors are relevant to consider in COVID-19^34,35,47^. As COVID-19 is a novel disease, we anticipate that controlling for other weather factors such as dew point, pressure, wind speed, and wind gust may increase the robustness of the conclusions. With this rationale, we derive twelve criteria for evaluating the relevant literature exploring the role of weather factors in COVID-19, as shown in Table 1.

**Table 1:**
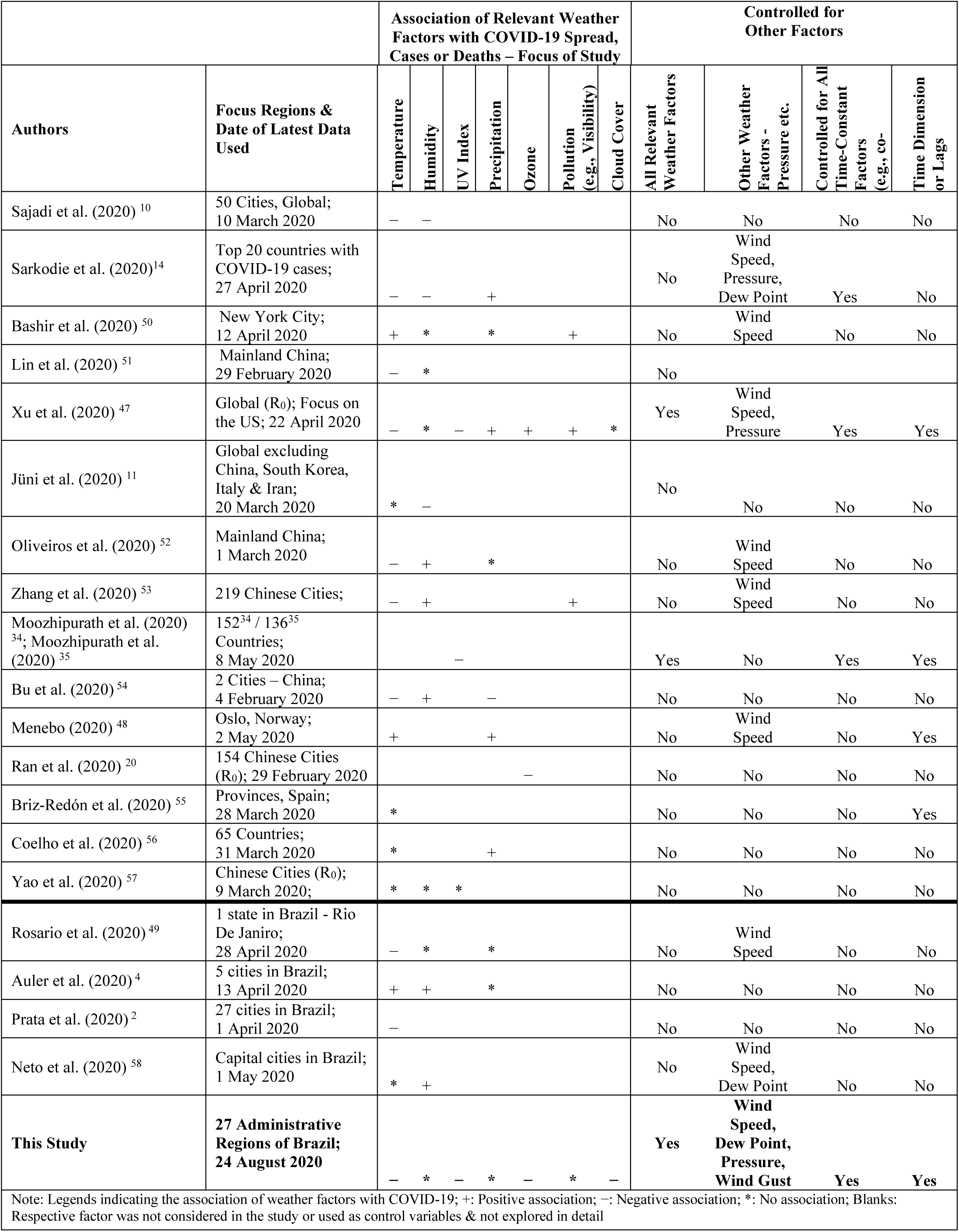
Review of Studies Exploring Weather Factors in COVID-19.

We provide an overview of the evaluation of various studies focusing on the role of the above relevant weather parameters in COVID-19, as shown in Table 1. Although COVID-19 studies indicate that these relevant weather factors may influence COVID-19 cases as well as deaths, the magnitude and direction of those associations remain inconclusive. Results of some studies a indicate that COVID-19 spread and plausibly deaths associate negatively with temperature^10,15,47,49–51^, humidity^10,15,52,53^, UV Index^34,35^, precipitation^48,54^, ozone^20^. However, the results of other studies indicate no association with temperature^55,56^, humidity^56^,precipitation^50^, UV radiation^57^. Some studies even indicate a positive association with temperature^48,52^, precipitation^15,47^, pollution^53^ and humidity^53^.

Further, studies exploring the impact of these weather factors on a tropical country like Brazil are limited. A few studies focused on Brazil find a negative association with temperature^2,49^; whereas others show a positive association with temperature^4^ and humidity^4,58^ or no association with temperature^58^. Further, as shown in Table 1, the majority of the Brazilian studies used data for a shorter period covering only the time until the start of Brazilian winter. Furthermore, no studies in Brazil has so far considered weather factors such as UV Index, ozone, visibility and cloud cover.

The reasons for the above inconclusive results are unclear^47^. As mentioned in Table 1, most of these studies considered only a subset of these weather factors. They did not take into account the lags, which might potentially limit the reliability of the results^47^. Further, the majority of these studies did not take into account the above time constant confounding factors. Other limitations include using Reproduction Number (R_0_) of COVID-19 as the dependent variable, whose estimation includes several assumptions^47^. Majority of the studies had over-representation of locations with a lower temperature or lower UV^47^ either due to various studies conducted during northern hemisphere winter/spring or due to the focus on regions in the higher latitudes in the northern hemisphere.

Extending the study by Moozhipurath, Kraft, and Skiera (2020) ^34^ and Moozhipurath, and Kraft (2020) ^35^, we investigate the role of relevant weather factors in COVID-19, focusing on the within-country variation in a tropical country, Brazil. In contrast to Moozhipurath, Kraft, and Skiera (2020) ^34^ and Moozhipurath, and Kraft (2020) ^35^ which focus on UVI, we evaluate the existing literature on all relevant weather factors in COVID-19 in detail. We further empirically explore the role of all relevant weather factors like temperature, humidity, UVI, precipitation, ozone and pollution in COVID-19 with a focus on Brazil. Furthermore, Moozhipurath, Kraft, and Skiera (2020) ^34^ and Moozhipurath, and Kraft (2020) ^35^ studied COVID-19 when Northern Hemisphere transitioned from winter to summer with the majority of the outbreaks focused on countries in higher latitudes. These countries in higher latitudes undergo substantial variation in temperature and UVI ^4^. It is not yet clear whether the results, i.e., the protective role of UVB radiation in COVID-19 is valid for a tropical country like Brazil, with higher UVI and limited variation in UVI and temperature. Contrary to the above studies, we also use a more comprehensive weather dataset focusing on Brazil, a tropical country in Southern Hemisphere with additional weather factors such as wind speed, dew point, pressure and wind gust. We also use a more extended period of data.

In this observational study, we empirically describe the role of these seven relevant weather factors in COVID-19 deaths in Brazil. Contrary to the majority of other studies, we focus on the within-country variation located primarily in the Southern Hemisphere tropical region, consider time lags and control for all time-constant and various time-varying confounding factors. Further, we consider all relevant weather factors as well as analyze extended periods of data. In robustness checks, we also incorporate additional time-varying weather parameters such as dewpoint, pressure, wind speed and wind gust to ensure the robustness of our results.

## 4 Methods

### 4.1 Description of Data

We sourced COVID-19 data, covering across 27 administrative regions (26 states, 1 Federal District) of Brazil from the publicly available data published by the Ministry of Health, Brazil (https://covid.saude.gov.br/). The dataset covers 182 days from 25 February 2020 until 24 August 2020. We then used Python Geocoder to identify the latitude and longitude of these 27 administrative regions of Brazil (26 states, 1 Federal District). We then collected the daily weather data from https://darksky.net/ for these 27 administrative regions, based on the above latitude and longitude information. Subsequently, we merged these data to construct the final dataset at an administrative region level, as outlined in Table 2. All of these 27 administrative regions have reported more than 20 COVID-19 cumulative infections as of 24 August 2020. To ensure that the observations at the early stage of COVID-19 do not bias our results, we removed all 20 daily observations at the beginning of the outbreak for each administrative region.

**Table 2:** Summary of the Dataset.

|  |  |
| --- | --- |
| Number of administrative regions of Brazil in our data set | 27 regions<br>(26 states, 1 Federal District) |
| ... > 0 cumulative COVID-19 deaths before 24 Aug 2020 | 27 regions<br>(26 states, 1 Federal District) |
| ... > 20 cumulative COVID-19 infections before 24 Aug 2020 | 27 regions<br>(26 states, 1 Federal District) |
| Time-period | 25 February 2020 – 24 August 2020<br>(Duration: 182 days) |
| Level of data granularity | Daily |
| Data Source of daily COVID-19 deaths & cases at an administrative region level | Ministry of Health, Brazil,<br>https://covid.saude.gov.br/ |
| Latitude and longitude information for each administrative region | Python Geocoder |
| Weather data (daily) | Dark Sky<br>https://darksky.net/ |

The dataset consists of the cumulative daily number of COVID-19 deaths at an administrative region level along with relevant weather factors such as temperature (minimum and maximum – both in degrees Celsius (°C)), relative humidity (percentage, scale 0% - 100%)), UV Index (on a linear scale : 0 - 11+)), precipitation (mm), ozone (Dobson Unit (DU)), visibility (Kilometers (KM)) and cloud cover (percentage of cloud cover, on a scale : 0 - 1). UV Index is measured at ground level. Table 3 summarizes the descriptive statics of our dataset. As of 24 August 2020, on average per 27 administrative regions in Brazil, cumulative COVID-19 deaths were 4,270, and the growth rate of daily COVID-19 deaths was 0.007. The average growth rate of daily COVID-19 deaths was 0.05 across administrative regions, and the study period. Minimum temperature and maximum temperature across these administrative regions and the study period were 17.9 °C and 29.0 °C respectively. Further, relative humidity and UVI were on average, 75.9% and 7.4, respectively. Precipitation, ozone, visibility and cloud cover were on average 0.14 mm, 15.7 KM, 257 DU and 0.45 or 45% respectively.

**Table 3:**
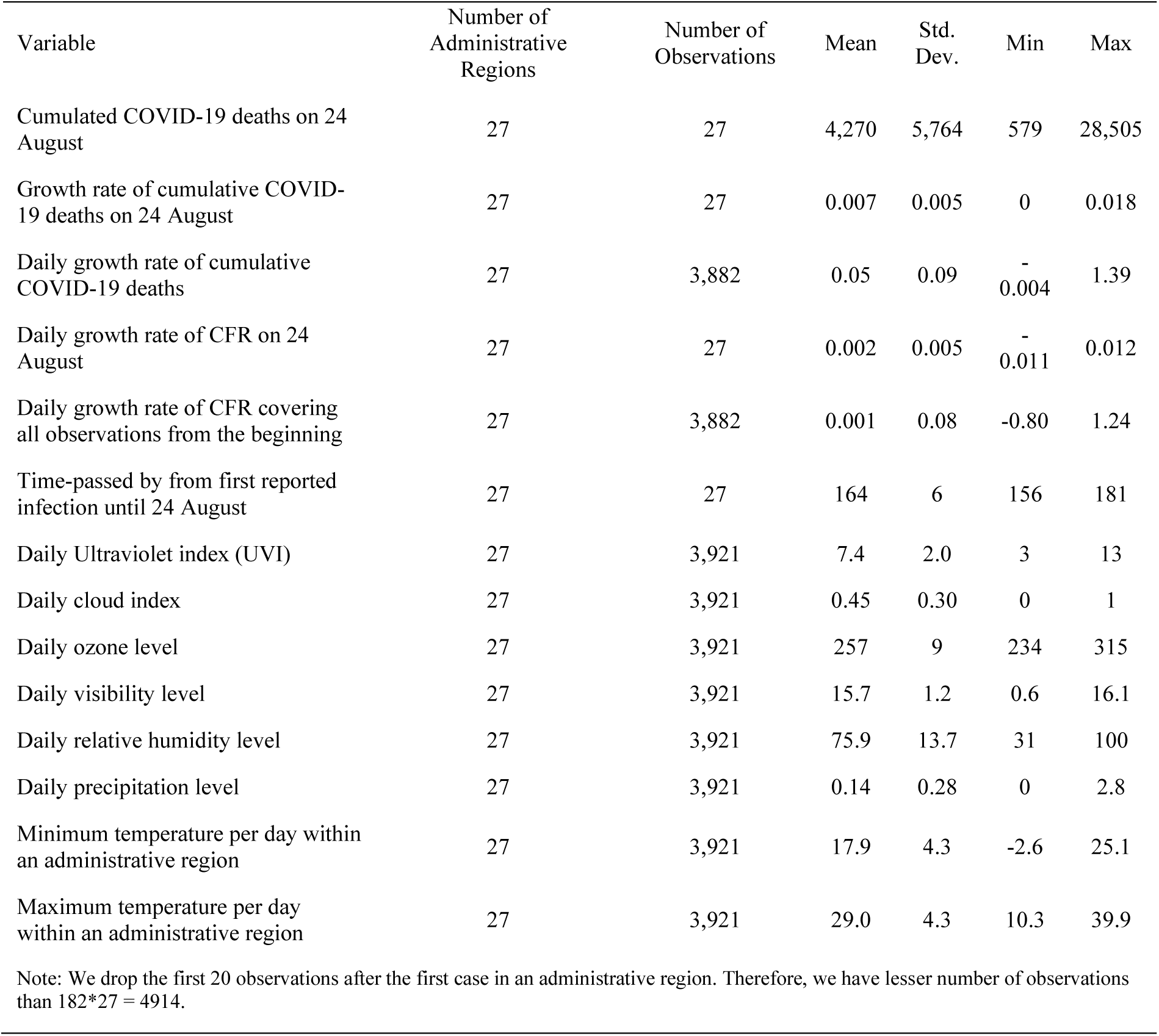
Descriptive Statistics of Data Set.

### 4.2 Description of Methodology

We use a log-linear fixed-effects model to the above panel dataset of 27 administrative regions in Brazil across 182 days (n=3882). We use the cumulative COVID-19 deaths as the dependent variable and the case-fatality-rate (CFR) for robustness checks. Further, our model isolates the association of these relevant weather factors from all time-constant and various time-varying region-specific factors. We account for the lagged effects of these weather factors by including 42 days moving averages of these weather parameters daily per region. Our fixed-effects model demeans the structural model and isolates these weather factors from region-specific time-constant factors analytically.

Our fixed-effects control for all region-specific time-constant factors such as age structure (e.g., fraction of population above 65 years of age), genetic profile, culture, density, age structure, gender, underlying disease, co-morbidities, skin pigmentation, gender, mobility, underlying conditions, diet or socio-economic factors that are relevant to consider in COVID-19^34,35,47^ We provide a detailed explanation of our methodology and the interpretation of these associations in section 1 and section 2 in the *Supplementary Appendix*.

## 5 Results

We summarize our main results in Table 4. After controlling for all time-constant and various time-varying confounding factors, we note a substantial and significant negative association of daily growth rates of cumulative COVID-19 deaths (COVID-19 daily deaths growth rate) with UVI, cloud cover, maximum temperature and ozone, with a moving average window of 42 days (Model 1 in Table 4). Specifically, a permanent unit increase of UVI, maximum temperature and ozone are independently associated with a decline of 6.0 percentage points [p<0.001], 1.8 percentage points [p<0.01] and 0.3 percentage points [p<0. 1] in COVID-19 deaths growth rate. A unit percentage increase in cloud cover is associated with 0.148 percentage points [p<0.05] decline in COVID-19 deaths growth rate. These results indicate their respective protective roles.

**Table 4:**
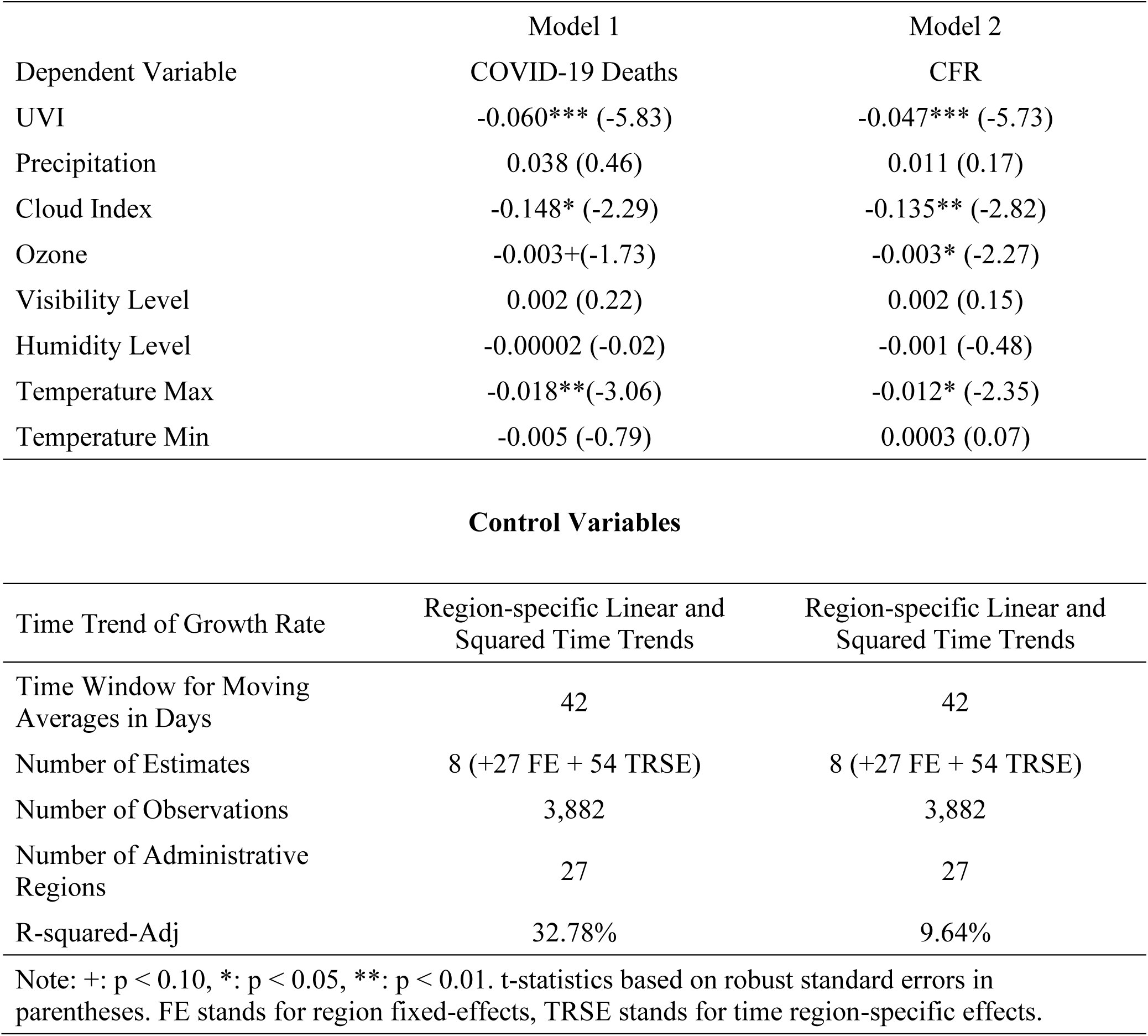
Effect of Weather Factors on Cumulative COVID-19 Deaths in Brazil.

Further, we explore what these associations mean in relative terms over a period of 2 weeks. A unit increase of UVI, maximum temperature and ozone are associated with a decline of 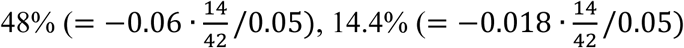, and 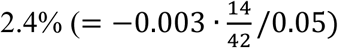 of daily growth rates of COVID-19 deaths as compared to the average daily growth rates over two weeks (14 days). As these results indicate, the above associations are substantial in relative terms, over a period of two weeks. Model 2 of Table 4 outlines that the results are consistent, even when we use CFR as the dependent variable. The estimated associations in Model 2 are weaker than those of Model 1 as the weather factors not only affect the COVID-19 deaths but also the infections, as outlined in section 2 of this manuscript.

We check the robustness of our results in Table S1 in *Supplementary Appendix*. We include additional time-varying weather parameters such as dewpoint, pressure, wind speed and wind gust. We find our results to be consistent across various model specifications, especially for UVI and cloud cover, even after incorporating these additional time-varying weather parameters.

## 6 Discussion

We observe a significant negative association of COVID-19 daily death growth rates in Brazil with weather factors - UVI, temperature, ozone and cloud cover. Specifically, a unit increase in UVI, maximum temperature, and ozone independently associate with 6.0 percentage points [p<0.001], 1.8 percentage points [p<0.01] and 0.3 percentage points [p<0. 1] decline in COVID-19 daily deaths growth rate. Further, a unit percentage increase in cloud cover associates with a decline of 0.148 percentage points [p<0.05] in COVID-19 daily deaths growth rate. These results translate into a substantial decline of COVID-19 daily deaths growth rate as compared to the average daily growth rates over two weeks (14 days) in relative terms especially for UVI (48%) and maximum temperature (14.4%). These results assume importance since the average UVI and average temperature are relatively high and their variations are moderate in tropical regions compared to other regions. Surprisingly, contrary to other studies, we do not find any association between COVID-19 daily deaths growth rate and humidity, visibility and precipitation. We find our results to be consistent even when we use the CFR as the dependent variable.

Our results are consistent with various theoretical mechanisms through which weather factors may affect immunity, human behaviour as well as viral survival and transmission^5^. Prior studies indicate that in higher latitudes (above 35° latitude) during winter, ozone absorbs most of the UVB^59^ and reduces the protective role of UVB. However, it is surprising that even in a tropical country like Brazil with high average UV Index and most of the geographic area below 33°S, UVB radiation plays a crucial mitigating role in COVID-19. These results are consistent with the emerging observational and clinical findings related to the protective role of UVB radiation due to its role in the inactivation of viruses as well as vitamin D synthesis^34,35,36,37,38^. This finding can guide the policy decisions, for example – sensible sunlight exposure and vitamin D supplementation – even in tropical countries.

We anticipate that temperature may play a protective role in COVID-19 in Brazil since higher temperature reduces the spread of COVID-19 and thereby the likelihood of deaths plausibly due to its effect on viral survival and transmission. These results are consistent with the majority of the existing studies on other viruses such as SARS-CoV^60^. In contrast with some prior studies, we find a protective role of ozone in COVID-19^20^ in Brazil^47^. This protective role may be plausibly due to its anti-viral effects^20^. Finally, the protective role of cloud cover in Brazil may be due to the changes in the behaviour of people – for example – more time spent indoors and thereby reducing the likelihood of transmission of COVID-19 outdoors. In contrast to many other studies^47,10,15,52,53,48,54^, we do not find any association between COVID-19 daily deaths growth rate and other weather factors such as humidity, visibility and precipitation in Brazil. We observe our results to be consistent across various model specifications, especially for UVI and cloud cover, even after incorporating additional time-varying weather parameters such as dewpoint, pressure, wind speed and wind gust.

We control for all time-constant and region-specific factors and different time-varying confounding factors^34,35^. Nevertheless, we admit that we may not be able to exclude other time-varying confounding factors, which might affect our results^34,35^. We also acknowledge that our study does not take into account the regional variations within Brazil as well as the difference in UVB exposure across urban and rural regions due to different amount of time spent outdoors. Our study uses the latest COVID-19 data with one of the comprehensive datasets available on weather in Brazil at an administrative region level.

In general, tropical countries do not undergo substantial variation in temperature and UVB radiation across seasons. However, our results indicate that even in such tropical countries, a consistent drop in UVI or temperature due to change of seasons (e.g., monsoon, rainy season, winter) may be associated with a substantial increase in the daily death growth rate of COVID-19. These results also serve as an early warning for tropical regions in the Northern Hemisphere which is transitioning to autumn and winter. Furthermore, our study indicates that with the arrival of summer (higher temperature and higher UVI), tropical countries in the southern hemisphere like Brazil may face a decline in COVID-19 deaths. We further acknowledge that our study results cannot be a substitute for health guidance for Brazil. However, we hope that the results of our study will guide further ecological studies and guide policy decision making in Brazil and other similar tropical countries.

## Data Availability

The data used in the study are sourced from publicly available sources. Data regarding COVID-19 are obtained on 25 August 2020 from the data released by the Ministry of Health, Brazil and can be accessed at https://covid.saude.gov.br/. Interested researchers can contact Rahul Kalippurayil Moozhipurath via email to get access to the data.

## 7 Declaration of Interests

RKM is a PhD student at Goethe University, Frankfurt. He is a full-time employee of a multinational chemical company involved in vitamin D business and holds the shares of the company. This study is intended to contribute to the ongoing COVID-19 crisis and is not sponsored by his company. The views expressed in the paper are those of the authors and do not represent that of any organization. No other relationships or activities that could appear to have influenced the submitted work.

## 8 Acknowledgements

We would like to thank Lennart Kraft for his valuable support, providing inputs to this paper and his review. We would like to acknowledge Sharath Mandya Krishna, for his support to this paper. We thank Matthew Little for his inputs and his assistance in the review. We would also like to acknowledge Magdalena Ceklarz for her contributions to our paper and the discussions about COVID-19 at different points in time.

## 9 Author Contributions

RKM conceptualised the research idea, conducted literature research, designed theoretical framework and collected COVID-19 data. RKM analyzed and interpreted the results and wrote the article.

## 10 Role of the Funding Source

This study is not sponsored by any organisation. The corresponding author had full access to all the data and had final responsibility for the submission decision.

## 11 Additional Information

Correspondence and requests for materials should be addressed to Rahul Kalippurayil Moozhipurath.

## 12 Data Sharing

The data used in the study are sourced from publicly available sources. Data regarding COVID-19 are obtained on 25 August 2020 from *the data released by the Ministry of Health, Brazil* and can be accessed at https://covid.saude.gov.br/. Data regarding weather is obtained from *Dark Sky* on the 25 August, 2020 and can be accessed at https://darksky.net/. We will make specific data set used in this study available for any future research. Interested researchers can contact Rahul Kalippurayil Moozhipurath via email to get access to the data.

## Notes

### Summary of Updates

1. Revised section 6 based on the feedback 2. New supplementary materials are uploaded

