## Supplementary Material for "Role of Weather Factors in COVID-19 Death Growth Rates in Tropical Climate: A Data-Driven Study Focused on Brazil Manuscript"

### COVID-19 Death Growth Rates

### in Tropical Climate:

### A Data-Driven Study Focused on Brazil

### Supplementary Appendix

Rahul Kalippurayil Moozhipurath*[1]

(Faculty of Economics and Business, Goethe University Frankfurt[1])

**Date:** 05.10.2020

Rahul Kalippurayil Moozhipurath, PhD Student; Faculty of Economics and Business, Goethe University Frankfurt, Theodor-W.-Adorno-Platz 4, 60629 Frankfurt, Germany;, Phone: +49-152-1301-0589;

### 1 Description of Methodology

In this supplementary section, we outline our methodology. We apply a Fixed-Effects log-linear regression model to estimate the effect of different weather parameters on the number of COVID-19 deaths. This fixed-effects model is closely related to the models proposed by Moozhipurath, Kraft, and Skiera (2020)[1], Moozhipurath, and Kraft (2020)[2] as well as Hsiang et al. (2020)[3]. This study extends the methodology proposed by Moozhipurath, Kraft, and Skiera (2020)[1], Moozhipurath, and Kraft (2020)[2] by exploring all relevant weather factors rather than only UV Index and also includes additional weather factors such as dewpoint, wind speed, wind gust and pressure. In contrast to Moozhipurath, Kraft, and Skiera (2020)[1], Moozhipurath, and Kraft (2020)[2] who use lagged values, we use a moving average of weather factors with varying time windows. A log-linear model increases the comparability of the growth rates of COVID-19 deaths across administrative regions because it considers percentage rather than the absolute changes over time[1,2]. Percentage growth rates are more comparable across administrative regions rather than the absolute ones[1,2].

Our model isolates the effect of weather parameters from region-specific time-constant factors via the fixed effects[1,2]. These time-constant factors consist of the location of the administrative region (as measured by its latitude and longitude) and other population factors such as demographics, age composition, gender, genetics, culture, location[1,2]. They also account for mobility & lifestyle of the population, the prevalence of co-morbidities, obesity and other chronic diseases and skin pigmentation[1,2]. Thus it controls for factors which may be associated with the severity of COVID-19[1,2]. These fixed-effects further capture time-constant diet-related factors such as the proportion of vegans and vegetarians, consumption of dietary supplements, fortified foods and diets rich in vitamin D, which might affect the growth rates of COVID-19 deaths[1,2]. Furthermore, they also capture the socio-economic situation of a population in an administrative region which are likely to remain reasonably stable over our observation period[1,2].

Further, the methodology also partially controls for the increasing pressure on the healthcare system in an administrative region over time[1,2]. We do so by flexibly controlling for the time passed by, since the first reported case of COVID-19 in each administrative region[1,2]. Importantly, this factor also helps to partial out any linear or quadratic change of growth rates over time that is similar across administrative regions[1,2]. Therefore, the model isolates the effect of the weather factors from the typically observed exponential-shaped or S-shaped curve, which are often found in the cumulative COVID-19 daily deaths over time[1,2].

The fixed-effects also capture time-constant behaviours of individuals such as recurring habits or mobility of individuals. These time-constant behaviours might affect the likelihood of exposure to weather factors (e.g., walking to work, outdoor exercises, outdoor activities related to employment) [1,2]. Yet, the fixed-effects do not capture time-varying behaviours of individuals such as their varying travel patterns and thus varying exposure to weather factors[1,2].

Because weather plausibly affects deaths several weeks later, we use the moving average of the previous 42 days of the weather factors in our model. The results did not change substantially, even after increasing or decreasing the time windows. We derive the following model in equation (1) to explain the number of COVID-19 deaths:

$$D_{i,t} = D_{i,t-1} \times e^{\gamma + W_{i,t}\beta_W + C_{i,t}\beta_C + u_i + \epsilon_{i,t}} \quad (1)$$

$D_{i,t}$ represents the cumulative COVID-19 deaths in the administrative region $i$ at time point $t$ (in days) . $D_{i,t}$ is related to the explanatory factors via an exponential growth model on the right-hand side of the equation in line with Moozhipurath, Kraft, and Skiera (2020)[1] and Moozhipurath, and Kraft (2020)[1,2]. The exponential growth model flexibly allows different shapes of the cumulative COVID-19 deaths[1,2]. The exponential growth model that we use in the study consists of five explanatory parts as shown below[1,2].

1) $\gamma$ represents the daily growth rate of COVID-19 deaths from $D_{i,t-1}$ to $D_{i,t}$ that is independent of the weather variables of this model. $\gamma$ controls for virus-specific attributes like its transmission characteristics such as basic reproductive rate $R_0$ combined with its lethality[1,2], similar to Moozhipurath, Kraft, and Skiera (2020)[1] and Moozhipurath, and Kraft (2020)[1,2].
2) $W_{i,t}$ represents the 42 days moving average of weather factors including ultraviolet index (UVI), precipitation, cloud index, ozone, visibility level, humidity level, the maximum and the minimum temperature for a state $i$ at day $t$. $\beta_W$ reflects the effect of weather factors.
3) $C_{i,t}$ stands for the set of control variables. In the main model specification, this set consists of the time passed by since the first reported COVID-19 infection for an administrative region $i$ at day $t$. In the specification for the robustness checks, this set also includes the moving average of additional weather variables. The vector $\beta_C$ represents the effect of the control variables in the equation.
4) $u_i$ represents time-constant region-specific factors influencing the growth rate of cumulative COVID-19 deaths (e.g., time-constant region-specific factors include population factors such as demographics, age composition, gender, genetics, culture, location, mobility, lifestyle, dietary pattern, dietary supplements, the prevalence of co-morbidities or other chronic diseases). The fixed effects isolate the weather factors from this time-constant state-specific factor, similar to Moozhipurath, Kraft, and Skiera (2020)[1].
5) $\epsilon_{i,t}$ consists of all the remaining factors which are not identified but also may have an effect on the cumulative daily COVID-19 deaths (i.e., all non-linear differences of

growth rates concerning time as well as the region-specific linear differences of growth rates concerning time), similar to Moozhipurath, Kraft, and Skiera (2020)[1]. These unidentified factors could be the declining number of people who could potentially become infected or contagious due to acquired immunity, lockdowns in an administrative region, mutation of the virus in an administrative region over time, systematic false-reports of the dependent variable by an administrative region[1,2].

An appropriate transformation of equation (1) results in the estimable equation (2) as given below.

$$\Delta \ln\left(D_{i,t}\right) = UVI_{i,t}\beta_{UVI} + C_{i,t}\beta_C + u_i + \gamma + \epsilon_{i,t} \tag{2}$$

If $\gamma$ and $u_i$ are correlated with past or future values of the weather factors, then the estimation of coefficients of equation (2) will be inconsistent. Therefore, we use a fixed-effects model that isolates the weather factors from those time-constant factors via the error correction term.

Equation (2) shows why we can only use those observations where cumulative COVID-19 deaths are greater than zero. We show an overview of how many observations per administrative region in Table S2.

### 2 Interpretation of Coefficients for Moving Average Variables

The structural model of equation (1) consists of 42 days moving average of the weather factors that are being studied. Therefore, the model outlines the impact of a consistent unit change in each of these weather factors (for example UVI) over this moving average window (e.g., 42 days) on the daily COVID-19 deaths growth rates after 42 days. We define this effect as the long-run effect. The linearity of the model implies that the short-run effect (e.g., the effect after 14 days) of the weather factors (e.g., UVI) on the growth rates of daily COVID-19 deaths equals the number of days of the considered period (e.g., 14 days) divided by 42 days (e.g., one third) times the long-run effect.

### 3 Identification of Effect of Weather Variables

Moozhipurath, Kraft, and Skiera (2020)[1]outlines the key assumption that is required to identify the causal effect of the UV Index on COVID-19 deaths. The key assumption required to identify the causal effect of the relevant weather factors is similar to that of Moozhipurath, Kraft, and Skiera (2020)[1], as UVI is one of the weather factors and has similar characteristics to the remaining weather factors. As such, a major threat to identification is the potential presence of a spurious correlation via time trends which could affect the weather factors as well as the growth rates of daily COVID-19 deaths. Therefore, we have to assume that all weather factors are uncorrelated to $\epsilon_{i,s}$ at all points in time conditional on the region-specific time-constant factors and the inclusion of the time trends of the model. Given the unavailability of variation of governmental measures across regions in Brazil, we are unable to control for such measures. However, because the weather factors are independent of the governmental measures, not controlling for governmental measures should not affect the consistency of the estimates.

### 4 Model Selection to Identify the Effect of Weather Variables on the Cumulative COVID-19 Deaths

We estimate equation (2) for different time windows of the moving averaged weather factors from 1 to 9 weeks. We do not observe any substantial changes concerning the size or the statistical significance of the estimates of weather factors.

### 5 Robustness Checks

Table S1 outlines the estimation with additional moving averaged weather factors such as dewpoint, pressure, wind speed and wind gust. We find consistent results across these model specifications as shown in Table S1. We also report the number of observations per administrative region and the latitude and longitude information which is used to collect the weather data per administrative region in Table S2.

### 6 Supplementary Tables

**Table S1: Effect of Weather Factors on Cumulative COVID-19 Deaths in Brazil (Robustness)**

| | Model 1 | Model 2 |
|---|---|---|
| Dependent Variable | COVID-19 Deaths | CFR |
| UVI | -0.062*** (-5.72) | -0.048*** (-5.14) |
| Precipitation | 0.029 (0.33) | 0.25 (0.34) |
| Cloud Index | -0.168* (-2.32) | -0.14* (-2.56) |
| Ozone | -0.0020 (-1.13) | -0.0028 (-1.70) |
| Visibility Level | -0.0012 (-0.11) | 0.00078 (0.07) |
| Humidity Level | (0.0027 (0.42) | -0.0024 (-0.39) |
| Temperature Max | -0.0079 (-0.61) | -0.014 (-1.19) |
| Temperature Min | -0.0018 (-0.11) | -0.006 (-0.36) |
| Dewpoint | -0.0065 (-0.23) | 0.011 (0.39) |
| Pressure | 0.0090+ (1.87) | 0.0025 (0.39) |
| Windspeed | -0.013 (-0.51) | 0.014 (0.55) |
| Windgust | 0.011 (1.46) | 0.000084 (0.01) |

**Control Variables**

| Time Trend of Growth Rate | Region-specific Linear and Squared Time Trends | Region-specific Linear and Squared Time Trends |
|---|---|---|
| Time Window for Moving Averages in Days | 42 | 42 |
| Number of Estimates | 12 (+27 FE + 54 TRSE) | 12 (+27 FE + 54 TRSE) |
| Number of Observations | 3,882 | 3,882 |
| Number of Administrative Regions | 27 | 27 |
| R-squared-Adj | 32.90% | 9.67% |

Note: +: $p < 0.10$, *: $p < 0.05$, **: $p < 0.01$. t-statistics based on robust standard errors in parentheses. FE stands for region fixed-effects, TRSE stands for time region-specific effects.

**Table S2: Number of Observations (Obs.), Latitude (Lat.) and Longitude (Long.) of Administrative Regions Used in Analysis**

| Region | Obs | Lat | Long | Region | Obs. | Lat | Long |
|---|---|---|---|---|---|---|---|
| AC Acre | 139 | -9.05 | -70.53 | PB Paraíba | 140 | -7.12 | -36.72 |
| AL Alagoas | 146 | -9.66 | -36.65 | PE Pernambuco | 147 | -8.4 | -37.59 |
| AM Amazonas | 144 | -4.48 | -63.52 | PI Piauí | 139 | -7.70 | -42.50 |
| AP Amapá | 139 | 1.35 | -51.92 | PR Paraná | 147 | -24.48 | -51.81 |
| BA Bahia | 148 | -12.29 | -41.93 | RJ Rio de Janeiro | 154 | -22.28 | -42.42 |
| CE Ceará | 142 | -5.33 | -39.72 | RN Rio Grande Do Norte | 146 | -5.68 | -36.48 |
| DF Distrito Federal | 148 | -15.78 | -47.80 | RO Rondônia | 139 | -10.94 | -62.83 |
| ES Espírito Santo | 144 | -19.57 | -40.17 | RR Roraima | 137 | 2.14 | -61.36 |
| GO Goiás | 146 | -15.93 | -50.14 | RS Rio Grande do Sul | 149 | -29.84 | -53.77 |
| MA Maranhão | 138 | -5.21 | -45.39 | SC Santa Catarina | 146 | -27.06 | -51.11 |
| MG Minas Gerais | 147 | -18.53 | -44.16 | SE Sergipe | 144 | -10.67 | -37.38 |
| MS Mato Grosso do Sul | 143 | -19.59 | -54.48 | SP São Paulo | 160 | -21.95 | -49.02 |
| MT Mato Grosso | 139 | -12.21 | -55.57 | TO Tocantins | 131 | -10.89 | -48.37 |
| PA Pará | 140 | -4.75 | -52.90 | | | | |
| Total Number of Observations | | | | 3,882 | | | |
